# Molecular and Microscopy Evaluation of Lymphatic filariasis in Ilorin Metropolis, Kwara State Nigeria

**DOI:** 10.1101/2025.01.03.25319939

**Authors:** Prosper Omah, Onyebiguwa Patrick Goddey Nmorsi, Andy Ogochukwu Egwunyenga

## Abstract

**Background:** Lymphatic filariasis is widespread and a major public health problem in many developing countries with warm and humid climate and it is one of the most prevalent neglected tropical diseases. This study, evaluated the diagnostics performance of molecular and conventional microscopy method of Lymphatic filariasis in Ilorin Metropolis, Kwara State Nigeria.

**Methods:** Genomic DNA was collected from 45 human blood samples obtained from Ilorin, Kwara State. They were then amplified by PCR and detected through gel electrophoresis. The PCR products were purified with QIAquick PCR purification kit (Qiagen, Valencia, CA) and then DNA quantified for Sanger sequencing, Bio Edit Software were used for sequence alignment and editing. 37 sequences were selected for analysis by MEGA X software to access the evolutionary connection of W. bancrofti isolates.

**Results:** 37 (82.2%) PCR products were positive for W. bancrofti and 16 (35.6%) were Positive for Microscopy. Hence, the sensitivity of PCR (82.2%) while the specificity was (64.4%).With the microscopy, the chances of a negative samples turning out positive by PCR, also known as the positive predictive value (PPV) was (69.8%) while the chances of being picked as truly negative predictive value was (78.4%). In assessing the level of agreement show that microscopy has a weak agreement, Kappa index statistics (0.46) with molecular techniques 95% CI,0.2-0.4).

**Conclusion:** Since Polymerase Chain Reaction showed higher sensitivity (82.2%) as positive predictive value, it is therefore recommended to include PCR as complementary routine diagnosis for lymphatic filariasis especially subclinical cases due to W. bancrofti despite its relatively higher cost.

## INTRODUCTION

Lymphatic filariasis (LF) is widespread and a major public health problem in many developing countries with warm and humid climate and it is one of the most prevalent neglected tropical diseases. *Wuchereria bancrofti* is the most prevalent cause of lymphatic filariasis cases in the world. The World Health Organization (WHO) considers lymphatic filariasis (LF) as the second leading cause of physical disability worldwide. An estimated 120 million people in 73 countries are currently infected, while 40 million people have chronic signs like hydrocele and elephantiasis [WHO,2016]. Lymphatic filariasis is widespread in sub-Saharan Africa and particularly high endemicity is seen along the coast of the Indian Ocean and in areas adjacent to the great lakes [Kwarleng,2019].Current estimates suggest that more than one billion people live in endemic areas and are at risk of infection, and more than one third of these are in Sub Saharan Africa [Christopher,2019].

Lymphatic filariasis, resulting from infection with the mosquito-borne filarial nematode *Wuchereria bancrofti*, is a disfiguring disabling disease [Kariuku *et al*,2011]. The parasites are transmitted from one host to another through the bites of infected mosquitoes which could be of different genera; *Culex, Aedes, Mansonia* and *Anopheles* [Hamalatha, *et al* 2016;Krol *et al* 2013 & Biritwum, *et al* 2016].The disorderly growth of the metropolis, the significant increase in the number of substandard housing and the parlous sewage disposal conditions favors proliferation of the vector mosquitoes and consequently transmission of the endemic diseases in the Africa region [Lau *et al* 2020].Therefore an intervention model for the control of lymphatic filariasis should consider the social organization of urban space by identifying micro areas of risk and priority population groups at risk of acquiring the parasites [Lau *et al* 2020;Rainima-Qaniuci *et al*, 2022]. Lymphatic filariasisis has thus been ranked the second most common cause of long term disability after mental illness [WHO,2017].Lymphatic filariasisis results in loss of work, productivity, direct and indirect economic loss and functional impairment [Okorie *et al* 2013;Kumar *et al*, 2016 & Stanton 2017],In Africa, the disease is endemic in 34 countries [Nosrat, 2021].

Nigeria has the greatest number of people infected or at risk with lymphatic filariasis (80-121 million), among all the African nations [Kariuku *et al*,2011;Ugboimoko *et al*,2010]. Lymphatic Filariasis affect many communities in Ilorin metropolis, neighboring villages especially local farmers causing illness and disfiguring that prevent people from attendance at school and farming. [Krol *et al,2013*].Most of the current diagnosis is based on the parasitological microscopy gold standard. Subclinical cases may be misdiagnosed using this techniques. This study is aimed to evaluate and compare the diagnostic performance of parasitological techniques and molecular technique using Real TimePolymerase Chain Reaction, (RT-PCR) for Lymphatic filariasis in our study area.

## MATERIALS AND METHODS

### Study Area and Design

The study was conducted in Ilorin, Kwara State. Ilorin is located approximately on latitude 8º30’ and 8º50’ North of the equator and longitude 4º20’ and 4º35’East of the Greenwich meridian. Ilorin is the gateway city between the southern and northern Nigeria with an approximate land area of 100kilometres .Ilorin has wet and dry season.The climate of the city is humid tropical under the influence of the two trade winds prevailing over the country, characterized with high temperature throughout the year .[Tamura *et al*,2013]. The daily average temperatures range change between September 22.5º to January 25 ºC. The mean annual rainfall is 1,200mm. The mean annual total rainfall is 1200m’The temperature in Ilorin is uniformly high throughout the year.. Ilorin metropolis is currently about 974,000 ranking 6th largest city in Nigeria. The climate of Ilorin is characterized by both wet and dry seasons. The rainy season begins towards the end of April and last till October while the dry season begins in November and end in April. The pattern of drainage system of Ilorin is dendritic. Asa River occupies a fairly valley and goes a long way to divide Ilorin into two parts: namely the Eastern and Western part. The wet season is between May and October with two peak periods in June and September while the dry season spans between November and April.

### Ethical Consideration

The ethical approved was obtained from Kwara State Ministry of Health Ethical Committee (MOH/KS//EU//777/354) before the commencement of the study The sample collection meeting was held with traditional leaders, teachers and communities members. Inform written consent was obtained from the volunteers, each adult subject and parents/guardian of each child during the study following the objectives of the study.Inventory of the volunteers were recorded for the study Right to refuse and withdraw were explained to the participant. The major inclusion criteria were presence of rashes,lymphedema and fever

### Study Subject And Population

The subject were 45 blood samples for molecular and microscopy comparisons The study was carried out between July, 2022 and August 2023 which spanned throughout the dry and rainy season.

### Sampling Procedure And Examination

#### Laboratory Analyses

##### Microscopy Examination

Whole blood samples were obtained by vein puncture at mid night and were submitted for microscopy in which laboratory DNA extraction, RT-PCR and sequences analysis laboratory blood filtration and microscopic evaluation of lymphatic filariasis due to *W*.*bancrofti* were performed by using 1ml of coagulated blood as described previously [Martindala *et al*,2024] using thick blood film why Meyer’s haematoxylin was examined microscopically and the level of microfilaeria documented [Nosrat,2021].

### DNA Extraction from human blood samples

DNA from human blood samples was extracted using the alcohol precipitation 200μl of blood was added in to labeled 1.5ml eppendorf tubes and 400μl of sodium hydroxide with 1% triton was added. The mixture was heated at 65°Cfor 30 minutes in a Thermomixer (Thermo Fisher) with continues shaking and quickly cooled on ice. Using a PH meter, the pH was adjusted to 8 for optimal performance of the Taq Polymerase during amplification. The DNA was centrifuged at 1400rpm, 4°C for 5 minutes and the supernatant transferred to a new eppendorf tube. Eight hundred (800) μl of absolute ethanol (98%) was added, vortexed and incubated overnight at - 80oC. The DNA was then washed thrice with 70% alcohol, dried and 50μl Tris-EDTA (TE) buffer added and stored at -20°c until amplification was performed.

### Detection of *W. bancrofti Using PCR*

PCR assays for detection of W. bancrofti were performed based on the sequence of the SspI repeat a pair of primers (NV-1andNV2) were designed or PCR amplification of the SspI repeat The target primers to amplify the DNA repeat was as follows for forward and reverse primers:NVi:5CGTGATGGCATCAAAGTAGCG3’;NV2:5CCCTCACTTACCATAAGACAAC3 [Martindala *et al*.2014]. The PCR products after amplification and detection by gel electrophoresis of *W. bancrofti* product size was 188bp against 100bp Molecular Ladder for the SspI repeat sequence.

### DNA purification and sequencing

After identifying positive PCR product samples (samples with the band of interest) from gel electrophoresis, the PCR products were purified with QIAquick PCR purification kit (Qiagen, Valencia, CA). After purification, gel electrophoresis was performed to check if the PCR products were present after purification. Sequencing PCR was performed in a Thermo Cycler using the purified amplicons, a single sequencing primer and a sequencing master mix The BigDye® Terminator v3.1 Cycle Sequencing Kits (Applied Bio systems, Foster City, CA). The sequencing protocol included one cycle of one minute at 95 °C followed with 50 cycles of 10 seconds of denaturation at 96 ° C, 5 seconds of annealing at 50 °C and 4 minutes of elongation at 60 °C and then a holding cycle at 4 °C. Then the Sequencing products were purified using Centri-SepTM spin columns (Princeton Separations, Inc New Jersey, USA). Followed by separation on a capillary ABI Prism 3 500 Genetic Analyzer (Applied Biosystems, Foster city, CA). The machine generates nucleotide sequence chromatograms that were obtained as ABI files format which were assembled and edited using sequence Scanner V1.0 (Applied Biosystems, Foster city, CA). Generated nucleotide sequences were edited by BioEdit® software and used in construction of a phylogenetic tree with the aid of MEGA 6.0® using the neighbor-joining method employing the Kimura-2 parameter [Kinyatta *et al*,2018;MOH-Kenya,2019]. Phylogenetic analysis of obtained *W. bancrofti Ssp 1* repeat sequences was 1 using the Kimura-2 parameter as implemented in MEGA 6.0. Phylogeny was inferred following 1000 bootstrap replication and reverse sequences were generated using the Bio Edit computer software

## PHYLOGENETIC ANALYSIS OF Ssp 1

MEGA X software was employed to access the evolutionary phylogenicity of *W. bancrofti* isolates from Ilorin metropolis. BLAST analysis on NCBI with the accession numbers LM12035, sequences of Ssp 1 gene in *W. bancroftil* using the technique developed by [MOH-Kenya,2019] The evolutionary history and distances between pairs of sequences were deduced using the neighbor joining methods algorithms. The branch of phylogenetic tree was statistically evaluated using a bootstrap of 1000 replication. BLAST was used to searched Genbank nucleotides data base homogeneous gene sequences to compare with the NCBI

## STATISTICAL ANALYSIS

Except sequencing all other data were analyzed using statistical package for social science (SPSS) version 21.0 (SPSS Inc. Chicago).The performance of each diagnostic test was calculated by means of specificity and sensitivity using positive predictive value and negative predictive value. To validate its measured by sensitivity and specificity they are best illustrated using a conventional two by two (2×2) table. MEGA X software was employed to access the evolutionary connection of *W. bancrofti* isolates Kappa’s statistics index was used to test the level of agreement between microscopic and PCR. P< 0.05 was considered statistical significant.

## RESULT

### PREVALENCE OF DIAGONISTIC PERFORMANCE OF PCR IN RELATION TO MICROSCOPY

Out of 45 blood sample examined for lymphatic filariasis due to *W. bancrofti* from the metropolis for molecular (PCR) and microscopy 37 (82.2%) were positive for PCR and 16 (35.6%) were Positive for Microscopy as shown in table1

**Table 1:** Comparison and agreement between result detected by PCR and microscopy.

| Diagnostic test | No Examine | Positive No(%) | Negative No(%) |
| --- | --- | --- | --- |
| PCR | 45 | 37 (82.2) | 8 (17.8) |
| Microscopy | 45 | 16 (35.6) | 29 (64.4) |
| total | 90 | 53(58.9) | 37 (41.1) |

**Table 2:** Evaluation Of Diagnostic Performance Of PCR Verses Microscopy.

| Diagnostic Test | PCR |  | Sensitivity(%) | Specificity (%) | PPV% | NPV% | Kappa's Index |
| --- | --- | --- | --- | --- | --- | --- | --- |
|  | Positive | Negative |  |  |  |  |  |
| Microscopy |  |  |  |  |  |  |  |
| Positive | 37 | 8 | 82.2 | 64.4 | 69.8 | 78.4 | 0.46 |
| Negative | 16 | 29 |  |  |  |  |  |

PPV = Positive Predictive value, NPV = Negative predictive value

### PCR SENSITIVITY AND SPECIFICITY

W .bancrofti isolates were subjected to MEGA X software to access their evolutionary phylogenicity relationship. The sensitivity of PCR was (82.2%) while the specificity was (64.4%).With the microscopy, the chances of a negative samples turning out positive by PCR, also known as the positive predictive value (PPV) was (69.8%) while the chances of being picked as truly negative predictive value was (78.4%). In assessing the level of agreement using microscopy as gold standard show that microscopy has a weak agreement, Kappa index statistics (0.46) with molecular techniques (PCR) (0.46, 95% CI,02-0.4).

### DNA Amplification Sequence Analysis

DNA was extracted from 45 human blood samples obtained from Ilorin metropolis They were then amplified by PCR and detected through gel electrophoresis. 37 PCR products positive for *W. bancrofti* were purified The PCR products after amplification and detection by gel electrophoresis are shown in figure Fig 2,3 and 4.The *W. bancrofti* product size was 188bp against 100bp Molecular Ladder for the SspI repeat sequence. Column A showed DNA marker ladder 100 bp, column B showed the sample 1 to 13, In Figure 2, B to N (sample Number 1-13) gave a positive result for DNA *W*.*bancrofti* at 188bp .In PCR Result of the sample 14 to 23 Column A showed DNA marker ladder 100 bp, column B to K showed the sample 1 to 11,column N was blank (No sample). In Figure 3, column B,C,D,E,F,,G,H,I,J K,L,and M (sample number 1-12) also gave positive results in 188bp while N was negative. In figure 4, Column A showed DNA marker ladder 100 bp, column B showed the sample 1 to 13, B to N (sample Number 1-13) gave a positive result for DNA *W*.*bancrofti* at 188bp. The PCR examination PPV was 69.8% and sensitivity was 82.2% while the NPV was 78.4% and specificity was 64.4%. A positive control (PC) known to be infected with *W. bancrofti* was used. After PCR, an expected 188 bp PCR fragment was produced in the PC and NC is negative control.

**FIG. 1:**
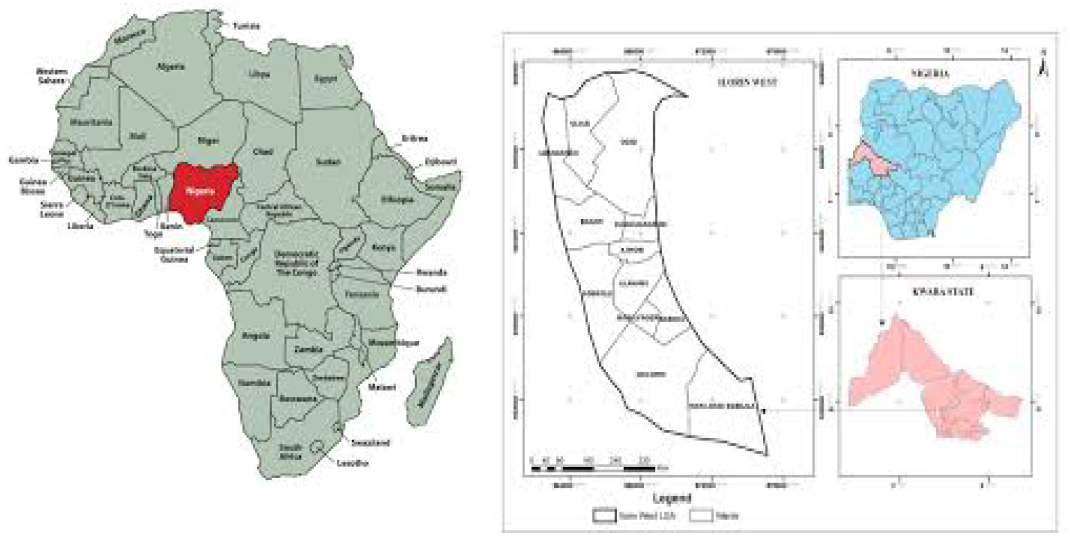
Map of Kwara State, showing the study area

**FIG 2:**
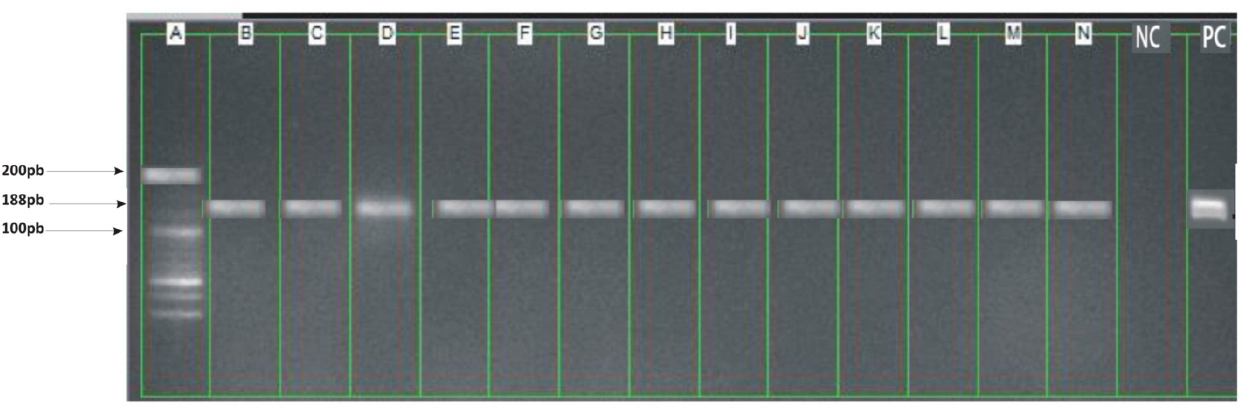
Detection of *W. bancrofti* DNA in Gel Electrophoresis using real time PCR.

**FIG 3:**
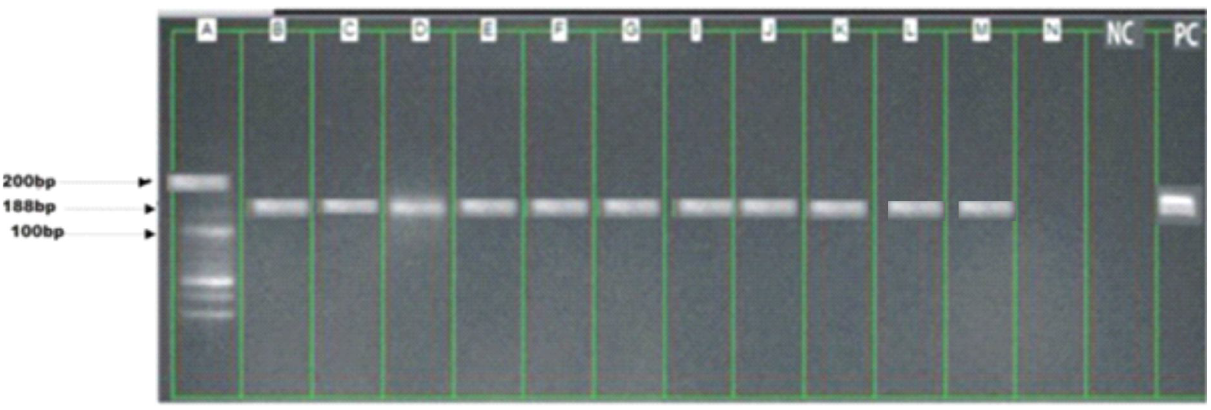
Detection of *W. bancrofti* DNA in Gel Electrophoresis using real time PCR.

**FIG 4:**
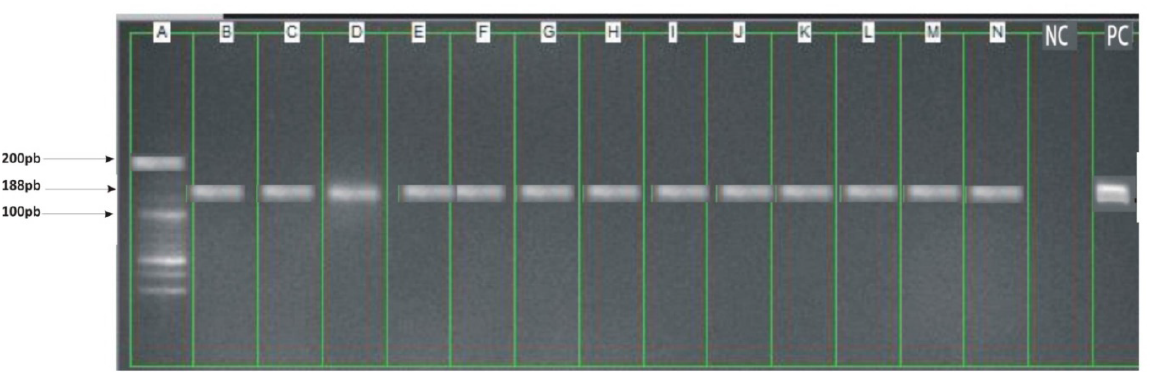
Detection of *W. bancrofti* DNA in Gel Electrophoresis using real time PCR.

### Phylogenetic Results

The phylogenetic analysis of SspI genes shows that the parasites were from three sites, the trees were drawn by neighbours joining method and supported with 1000 replicates bootstrap values which infer the phylogenetic trees. The division triangle, star and circle refers to the three types of parasitic variations within the species in Ilorin metropolis. Bootstrap value greater than or equal to 85% strongly support that the sequence data and branching order are accurate Fig 5. The evolutionary trees of the Ssp 1 genes in *W. bancrofti* was designed for the given sequences and the respective clades features were analyzed. The sequence Analysis of the amplicons of the samples investigated show a great number of similarities with W. bancrofti in Genbank according to BLAST analysis in NBCI .The genes were classified into three distinct clades based on the results of phylogenetic analysis and were indicated with symbols (circle, star and triangle) .Circle denotes samples from Ilorin West, star denotes Ilorin East and triangle denotes Ilorin South. Each clades was presented as a monophonic member of a given ancestor. Clades (star) 19 has the largest number of members followed by clades (circle) 10 which show a level of similarities in divergence from their respective ancestors while triangle 8 has the least number of members. In addition the *W. bancrofti* sequences in Ilorin West were not 100% identical but were closely related. *W. bancrofti* strain in some communities were not closely related. For instance Ilorin South but those in Ilorin East were 100% closely related.

**FIG 5:**
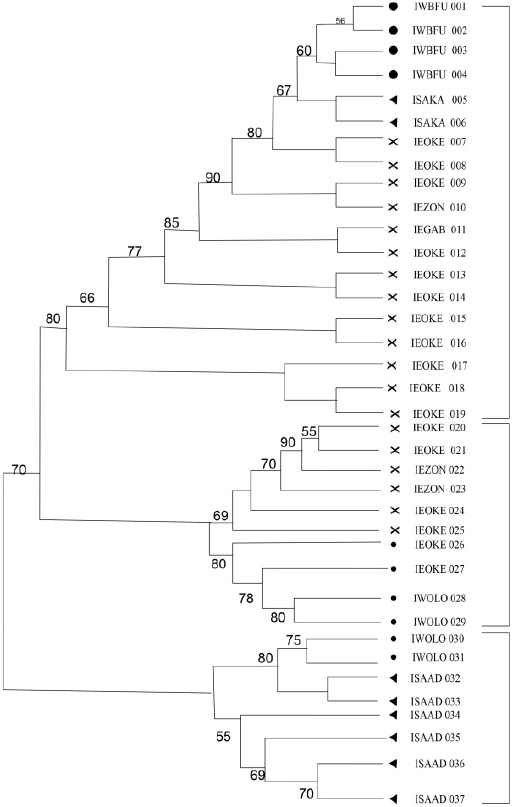
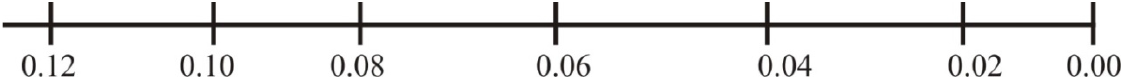
Evolutional Relationship of 37bp of Ssp1 gene of *W. bancrofti* using neighbouring-joining Phylogenetic Tree.

## DISCUSSION

The positive rate detected using PCR was 37(82.2%) is a strong evidence that detection of parasite specific DNA in blood is more sensitive and specific than processing blood by standard methods, microscopy [[Nosrat,2021;Njenga,2011;Ramash *et al*,2012]. These assays proved valuable tools for the diagnosis of lymphatic filariasis despite its relatively high cost as a diagnostic techniques. Our data corroborate with the finding of [Christopher,2019], who showed that PCR detected 22 positive cases among negative sample for blood filtration technique from individuals living in hypoendemic areas. Similar results were found [Biritwum,2016] using PCR and visualization on gel electrophoresis. [Kwarleng,2019] explained that the positive cases that were not detected using PCR could occur during the DNA amplification reaction as a result of various factors,. [De Souza *et al*,2012]. Sustained reduction in prevalence of lymphatic filariasis infection in spite of missed rounds of mass drug administration in an area under mosquito nets for malaria control..

The sensitivity and specificity of the SspI PCR system has demonstrated its potential use as a diagnostic tool. The result presented in this study demonstrated that the SspI PCR assay can be used to detect low numbers of parasite. However, sometimes microscopy examination gives a negative result on microfilaria especially when the presence were only in a small amount. A PCR can overcome this weakness and PCR examination using the parasite DNA have been reproduced into thousands of copies and give a positive outcome when the microscopic examination cannot detect small number of worms [Ramash *et al*,2012]. Study by [Nosrat,2021] demonstrated that PCR-ELISA assay analysis of blood sample from 206 individuals from an endemic area compared with samples slide from a single sample revealed a sensitivity of 9.74% and specificity of 85.1%. Which are lower than the values in our study [Christopher,2019] could be as a result of a recent population expansion. This difference may be explained by factors that are inherent in the experimental procedures, which may generate errors related to the detection of parasites DNA, its degradation during the biological sample collection or absence in the sample analysed. The need for a specific and accurate diagnostic test for LF are important, the current incidence of LF does not show any signs of decline, [Ramash *et al*,2012]. Without a sensitive and effective diagnostic test for the asymptomatic infections the accurate estimation of LF disease prevalence is not possible [Njenga,2011]. Without highly sensitive, specific tests, it will be difficult to ensure the objective control of LF. A recent publication from Thailand showed that filarial DNA could be detected by PCR analysis [Kariuku *et al*,2011]. However, the authors did not report results of specificity testing or results obtained with field samples. In agreement with results reported by [Ramash *et al*,2012] our preliminary studies showed that PCR work with NV1 and NV2 primers and SspI target sequence. The result suggests that DNA detected by PCR is more sensitive for detecting *W. bancrofti* DNA in field samples in endemic country laboratories [Ugbomoiko *et al*,2010] PCR can be applied to comparison between diagnostic methods and it is very useful to confirm suspicious cases that were not detected using conventional microscopy and evaluate cure control after chemotherapeutic treatment. This technique is important for long term success of parasite control by Mass Drug Administration (MDA) intervention because of the need to detect low intensity infections that persist even after treatment has occurred [Eward,2020]. This PCR technique is widely used for the diagnosis of several human diseases but its application in neglected diseases especially in Lymphatic filariasis was implored only recently. Some authors obtained high sensitivity and specificity [Okorie *et al*,2013] compared to routine microscopy.

The nucleotide amplification sequencing and phylogenetic construction of the SspI DNA repeat reveal that there are different strains of *W. bancrofti* circulating in the human blood samples in Ilorin Metropolis (Ilorin East, West and South). The findings show that *W. bancrofti* strains in some communities were not closely related, For instance *W. bancrofti* strain in Ilorin South and Ilorin West. *W. bancrofti* in the study area clustered which differ from the previously reported *W. bancrofti* in China using SspI repeat region .The current study revealed a considerable diversity between the parasite strain of the three places, the phylogenetic analysis exhibited three distinct clusters of *W. bancrofti* among Ilorin East, South and West populations. The existence of different strains of *W. bancrofti* was documented in the past [Dai, 2016] The parasite population of three locations of Ilorin metropolis exhibited very high genetic diversity formed at least three distinct clusters in the phylogenetic tree. Though few studies have pointed to genetic and morphological variation in *W. bancrofti* populations, our findings agree with a previous study by [Stanton, 2017], who reported the presence of two different genetic variants of the parasite with high genetic divergence and gene flow in different geo climate regions in India. The existence of considerable genetic variability of *W. bancrofti* using RAPD markers was obtained in southern parts of India [Stanton,2017][. Such variability has been reported for other human parasitic nematode such as *Ascaris* [Santon,2017] and *Trichuris trichiura* [Tamura *et al*,2013]. Hence, influx of inflected population will result in a mixing up of genetically different population and thereby a great genetic diversity. It may be noted that a patient filaria infection results from multiple infective bite to human population [Njenga, 2022]. Long term accumulation of infection brings genetic variants of the parasite in an infective individual. Thus increase in the genetic diversity and the genetic make-up of the parasite population in individuals may influence phylogenetic expressions in terms of drugs response, pathogenicity, clinical outcome and survival. The present study revealed the existence of genetically variant of parasite population of *W. bancrofti*. The study by [Biritwum,2016] showed that there is considerable genetic variability within *W. bancrofti* population in Ghana. The genes were classified into the district clades base on the results of the phylogenetic analysis. The presence of multiple identical members depicts their common ancestors origin and can be due to a recent gene duplication event [Rainima-Qaniuci *et al*,2022].

The data in this study shows that Ilorin South and Ilorin West samples are more distant. This study despite the small number of samples analyzed reveal some level of genetic variability in *W. bancrofti* in Ilorin metropolis thus, adding to the very few studies that aimed at understanding the genetic heterogeneity of *W. bancrofti* [Christopher,2019]. The genetic differences observed could be attributed to the environmental selection pressures. This may explain the epidemiological and field observations of lymphatic filariasis in Ilorin Metropolis. From the genetic tree the slightly closeness of samples from Ilorin East, South and Ilorin West could be as a result of migration of microfilaria individuals or vice versa. Migration is an important activity in Ilorin South and Ilorin West that occurs during specific times of the year, purposely for religious and economical activities could result in the transfer of parasites [Kumar *et al*, 2016;Martindale *et al*,2014]. These findings in the genetic heterogeneity of the population at different places within microfilariae carriers called for appropriate chemotherapy strategies for the elimination of lymphatic filariasiA [MOH-Kenya,2019] Furthermore, there have been reports of persistent residual Lf infections in some communities in Ghana and Burkina Faso, despite 5-8 rounds of treatments [Dai,2016;De Souza *et al* 2012], *Culex quinquefasciatus* strains from Liberia had a low susceptibility to *W. Bancrofti* from Liberia, while the same strain had a high susceptibility to *W. Bancrofti* from Sri Lanka. Consequently, the study concluded that the Liberian and Sri Lankan strains of *W. Bancrofti* differed in their ability to infect specific mosquito strains and therefore confirms our suggestion that the geographic strain of *W. bancrofti* can improve vector competence. The same reasons may explain the different epidemioogies of LF in East and West Africa, as to why *Culex quinquefasciatusis* not a vector of LF in West Africa, but is a very efficient in urban areas in East Africa [Kwarleng,2019]. However, this needs to be further investigated.In order to further explore the results produced in this study, we recommended that further studies with a much larger sample size and more detailed, population analyses be conducted on the diversity of *W. bancrofti* populations. This will enable the determination of profound conclusions on *W. Bancrofti* diversity. The availability of the complete mitochondrial genome sequence of *W. Bancrofti* [De Souza *et al*,2012;Kinyatta *et al*, 2018] will enhance a further understanding of phylogenetic and geographic relationships between isolates, and assessing population diversity within endemic regions.

## Conclusion

Molecular assay seems to be promising substitute for direct microscopy, Furthermore, another important aspect of molecular method is their applicability to epidemiological, such studies involved genetic diversity of populations and geographical distribution of parasitic diseases The analysis of genetic profiles of *Wuchereria bancrofti* from the three sites in Ilorin Metropolis indicated the existence of considerable genetic variability among parasites population in the study area. The study showed that *W*.*bancrofti* in Ilorin metropolis were highly variable.

## Data Availability

All data produced in the present study are available upon reasonable request to the authors

## Abbreviation

PCR: Polymerase chain reaction
DNA: Deoxyiribonucleic acid
LF: Lymphatic filariasis
USA: United states American
ELISA: Enzyme Linked Immuno absorbant assay
MDA: Mass drug administration
DALY: Disability adjusted life years
MOH: Ministry of health
WHO: World health organization
PPV: positive predictive value
NPV: Negative Predictive value

## Acknowledgement

The authors gratefully acknowledge the technical assistance of the National Institute of Medical Research Yaba Lagos, West Africa research Ibadan.

## Notes

### Competing Interest Statement

The authors have declared no competing interest.

### Funding Statement

This study did not receive any funding

### Author Declarations

The ethical approved was obtained from Kwara State Ministry of Health Ethical Committee (MOH/KS//EU//777/354) before the commencement of the study

